# Older Employees’ Perspectives on the wLiFE55+ Program: A Qualitative Evaluation of a Workplace Lifestyle-integrated Exercise Intervention

**DOI:** 10.1101/2024.12.19.24319321

**Authors:** Diana Pfister, Yvonne Ritter, Greta M. Steckhan, Susanne Voelter-Mahlknecht, Britta Weber, Rolf Ellegast, Markus Gruber, Michael Schwenk

## Abstract

**Background:** The ageing workforce and increasing prevalence of sedentary behaviour highlight the need for workplace strategies that promote physical activity (PA) and prevent functional decline in older employees. Traditional workplace PA interventions (WPAI) often lack feasibility due to limited time, low motivation, and insufficient integration into daily routines. The wLiFE55+ program applies a lifestyle-integrated approach by embedding strength, balance, and PA activities into everyday work tasks. This study qualitatively explored participants’ experiences to assess acceptability, perceived benefits, and priorities for refinement.

**Methods:** This qualitative sub-study was embedded in a four-week wLiFE55+ pilot intervention for employees aged 55 years and older. After completing the intervention, two focus groups (n = 8; mean age 58.5 years) were conducted. Discussions were audio-recorded, transcribed verbatim, and analysed using deductive qualitative content analysis informed by the Lifestyle-integrated Functional Exercise (LiFE) concept and the Theoretical Domains Framework. Analysis focused on three overarching categories: (1) overall program experiences, (2) perceptions of program content, and (3) behaviour-change processes.

**Results:** Participants perceived wLiFE55+ as meaningful, feasible, and aligned with naturally occurring work situations. Trainer support facilitated understanding, adaptation, and confidence in implementing activities. Reported challenges included insufficient preparatory information about the baseline assessment, rapid progression of activity difficulty, social discomfort in shared office environments, and the burden of paper-based documentation. Participants expressed a preference for clearer communication, more practice time between sessions, digital documentation and feedback tools, and an expanded set of activities, including options for the upper body.

**Conclusion:** Lifestyle-integrated approaches such as wLiFE55+ show promise for supporting functional capacity and promoting physical activity among older employees. Embedding activities into routine work tasks may help overcome common limitations of traditional workplace physical activity interventions. Future refinements should include improved preparatory guidance, greater environmental fit, and longer program durations to facilitate habit formation and sustainable implementation.

## Background

The ageing of the global workforce represents one of the most significant demographic and economic challenges of the coming decades [1]. As the proportion of employees aged 55 years and older continues to rise, organisations increasingly rely on retaining older workers to respond to labour shortages and maintain productivity [2–4]. Sustaining employability at older ages requires sufficient functional capacity. However, age-related declines in neuromotor function, muscular strength and aerobic capacity typically accelerate from about 55 years onwards [5–8]. These changes increase the risk of mobility limitations, chronic disease and reduced work ability [9, 10]. Although age-related physical decline most strongly affects physically demanding occupations, good physical functioning also supports work ability across all job types by reducing fatigue, enhancing safety and facilitating routine movement tasks. Sedentary behaviour, which is highly prevalent in office-based occupations, exacerbates this decline and may contribute to accelerated functional deterioration among older employees [11–15].

Current World Health Organization (WHO) guidelines recommend multicomponent physical activity (PA) for older adults, including strength and balance training, and emphasise that health benefits can accrue from small bouts of activity accumulated throughout the day [16]. Yet implementing these recommendations in real workplace settings remains challenging. Traditional workplace physical activity interventions (WPAI) often struggle with limited time availability, low relevance to employees’ daily tasks, workflow disruptions and inconsistent managerial support, resulting in modest adherence and limited behavioural change [17–23]. It is important to note that most existing evidence originates from high-income countries in the Global North, whereas findings from low-and middle-income countries highlight more diverse occupational PA patterns among older adults. In many low-and middle-income settings, older employees engage in more physically demanding work, whereas work in high-income countries is predominantly sedentary. These structural differences underscore the need for context-sensitive and adaptable approaches to workplace health promotion [24].

Lifestyle-integrated exercise approaches represent a promising alternative to traditional WPAI because they integrate functional movements directly into everyday routines rather than requiring additional training sessions. The Lifestyle-integrated Functional Exercise (LiFE) program exemplifies this concept by embedding strength and balance activities into daily tasks through personalised cues and contextual prompts [25]. Research across various LiFE adaptations—individual, group-based and technology-supported—shows high acceptability, strong adherence and improved functional outcomes among older adults [25, 26]. These findings suggest that embedding meaningful movements into familiar routines may reduce cognitive and motivational barriers, increase perceived relevance and enhance long-term adoption.

Building on this body of evidence, Ritter et al. developed wLiFE55+, a LiFE-based workplace intervention tailored to office-based employees aged 55 years and older. The program aims to integrate brief neuromotor, strength and PA tasks into commonly occurring work situations, while being feasible to implement within the constraints of a standard workday. A pilot feasibility trial demonstrated high adherence, improvements in neuromotor function and reductions in sedentary time [27]. However, quantitative findings alone cannot capture how employees experience the program in practice, how well activities fit into diverse workplace environments or which factors facilitate or hinder sustained engagement.

Qualitative inquiry is therefore essential to provide a more nuanced understanding of employees’ perspectives and implementation processes. Insights into perceived relevance, feasibility, behavioural mechanisms and contextual barriers can inform refinement of wLiFE55+ and support future scale-up. The present study explored older employees’ experiences with program content, delivery and behavioural integration following a four-week workplace intervention. We aimed to understand (1) overall program experiences and organisational context, (2) perceptions of specific program components and materials, and (3) behavioural processes related to motivation, planning and habit formation. These findings contribute to the optimisation of workplace lifestyle-integrated PA interventions and the development of sustainable strategies to promote functional capacity and active ageing in increasingly sedentary workforces.

## Methods

### Study design and setting

This qualitative study was embedded in the wLiFE55+ proof-of-concept pilot trial [27], which evaluated the feasibility of a four-week workplace lifestyle-integrated exercise program for employees aged 55 years and older. The four-week duration was intentionally chosen because the pilot trial aimed to assess feasibility, acceptability and implementation processes rather than to detect meaningful physiological changes, which would require a longer intervention period. The qualitative component was designed to complement the quantitative pilot findings by providing an in-depth understanding of participants’ experiences, perceived facilitators and barriers, and contextual factors influencing implementation.

A qualitative descriptive approach using focus group discussions was chosen to allow participants to collectively reflect on shared and divergent experiences. Two focus groups with four participants each were conducted after completion of the intervention and follow-up assessments. Small, homogeneous focus groups are recommended in feasibility research because they promote balanced participation and rich interaction [28, 29]. Sessions took place in seminar rooms at the University of Konstanz, lasted approximately 60 minutes, and were moderated by an independent researcher not involved in the intervention to minimise bias.

### Participants

Participants were recruited from the wLiFE55+ pilot trial, which included 15 employees aged 55 years or older. Inclusion criteria were current employment, age ≥55, and no contraindications for physical activity. A purposive sampling strategy ensured variation in working hours, physical activity and functional capacity. Eight participants (four women, four men; mean age 58.5 years, SD = 3.1) volunteered for the focus groups.

This sample size represents a typical proportion for embedded qualitative components in feasibility studies. Thematic saturation was reached, as no new themes emerged in the second focus group [30, 31]. All participants provided written informed consent.

### Program

wLiFE55+ integrates brief neuromotor, strength and physical activity tasks—such as one-leg stands, squats and brisk walking—into naturally occurring work situations. Table 1 provides an overview of activities; full program details are reported elsewhere [27].

**Table 1.**
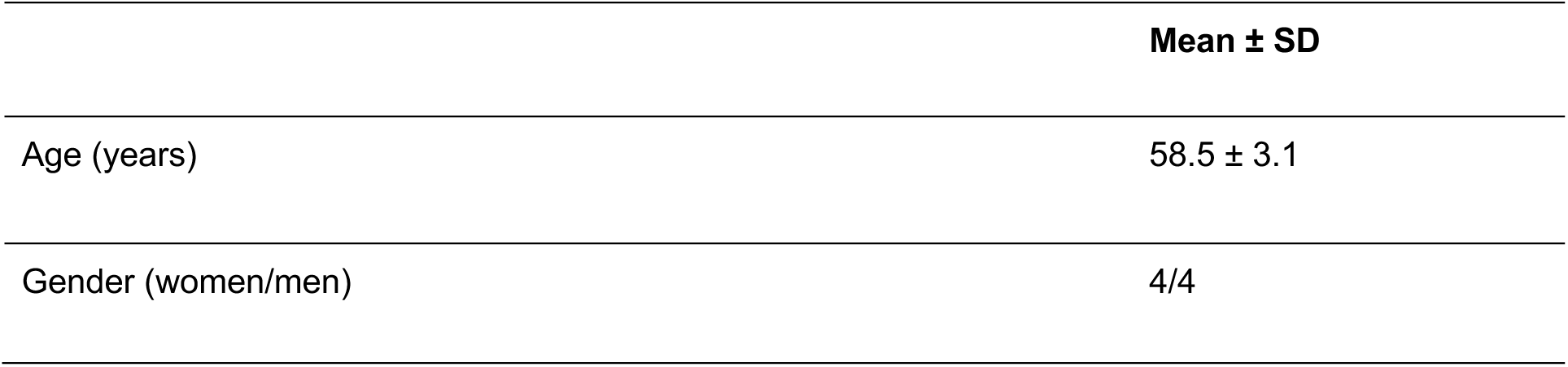
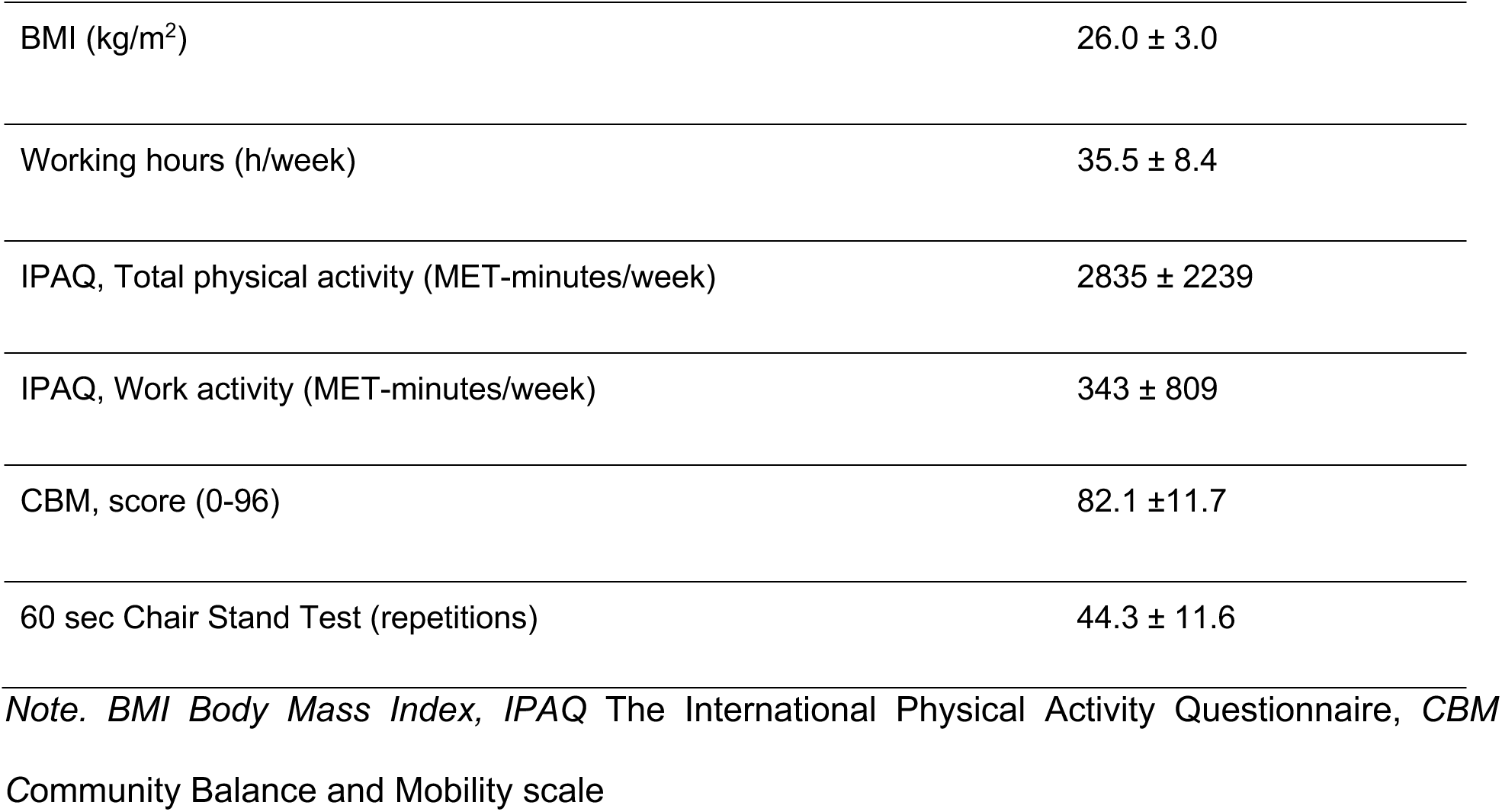
Characteristics of focus group participants (n = 8)

**Table 2.**
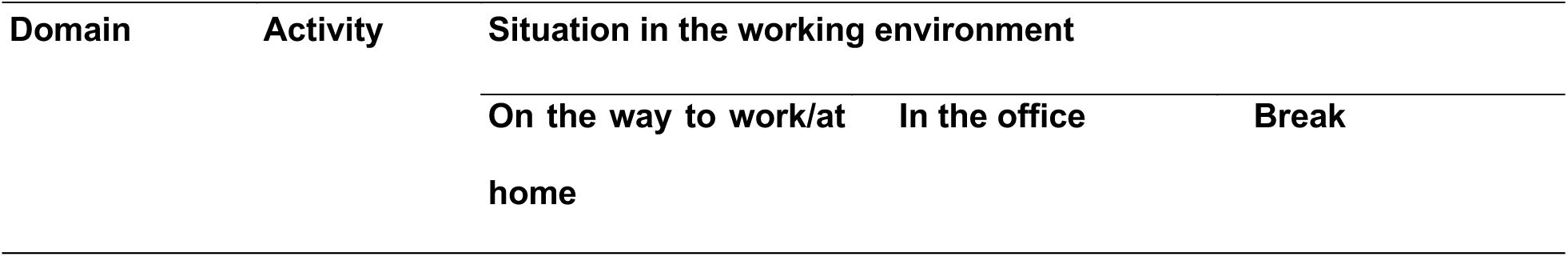
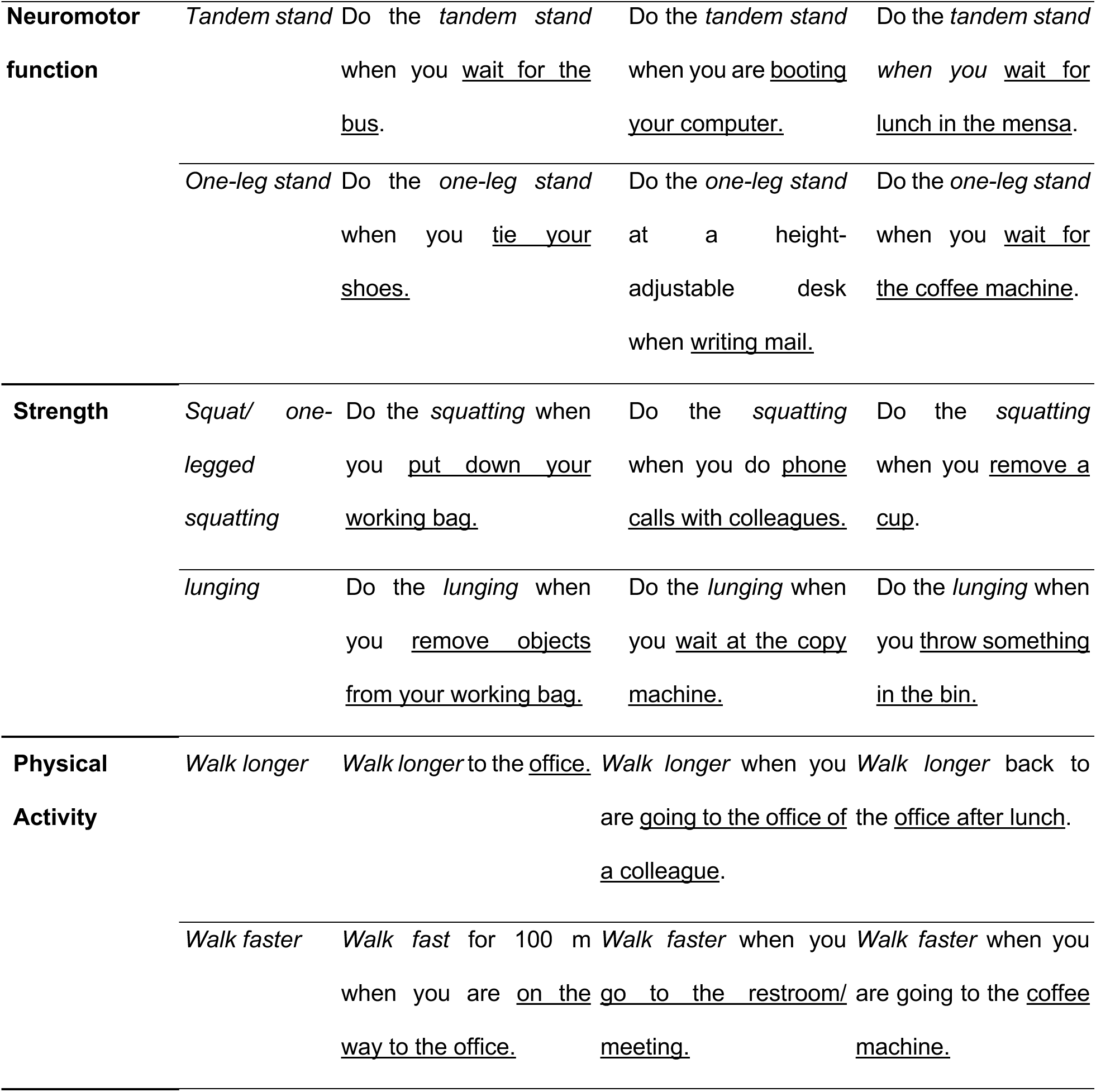
Short overview of wLiFE55+ activities with situations in the working environment (detailed table in [27])

Baseline and follow-up assessments were conducted either at the University of Konstanz or in the workplace to evaluate real-world feasibility. In the pilot trial, wLiFE55+ showed high feasibility, with 100% adherence and no adverse events. Quantitative pre–post analyses indicated medium-to-large improvements in neuromotor function (including significant CBM gains), reductions in sedentary time and sedentary bouts, and positive trends in lower-body strength and daily activity. As a feasibility study, the four-week duration was designed to assess acceptance and early implementation rather than fully developed physiological change [27].

These quantitative findings demonstrate feasibility and potential benefit, but they do not capture how participants experienced and implemented the program in daily work life—questions addressed by the present qualitative study.

Participants attended four weekly individual sessions with a trained personal trainer after the baseline assessment. Trainers introduced core activities, tailored difficulty levels, and supported participants in linking activities to daily work routines using planning tools described in the program manual. These sessions aimed to enhance understanding, personalize activities and support early habit formation.

### Administering the intervention

After the baseline assessment, participants received the wLiFE55+ manual and completed four weekly individual sessions with a trained personal trainer at their workplace. Trainers introduced core activities, assessed individual abilities and suitable difficulty levels, and supported participants in linking strength, balance and physical activity tasks to everyday work situations using tools such as the Daily Routine Chart and Activity Planner. Across sessions, trainers reviewed previously introduced activities, adjusted difficulty, provided feedback and refined cue–activity plans to support integration into daily routines. Documentation in the Activity Planner supported action planning in line with the LiFE concept and the Health Action Process Approach (HAPA). These sessions aimed to personalise activities, enhance capability and confidence, and facilitate early habit formation [27].

### Data collection

An interdisciplinary team of researchers from sports science (DP, YR, MG, MS), occupational health (GS, SVM), occupational safety (BW, RE) and the occupational healthcare industry (CK, FB) developed a semi-structured interview guide (Table 3) based on previous qualitative evaluations of LiFE [3–5]. Two focus groups (n = 4 each) were conducted by an independent moderator (MS) who was not involved in the intervention to minimise bias. Participants were encouraged to discuss their experiences openly and to include both positive and critical reflections. All discussions were audio-recorded and transcribed verbatim with a semantic focus following Kuckartz’s guidelines [32]. As the analysis targeted manifest content, non-verbal cues were not coded. The moderator actively encouraged balanced participation and monitored for dominance effects, which helped manage group dynamics in the small focus groups.

**Table 3.**
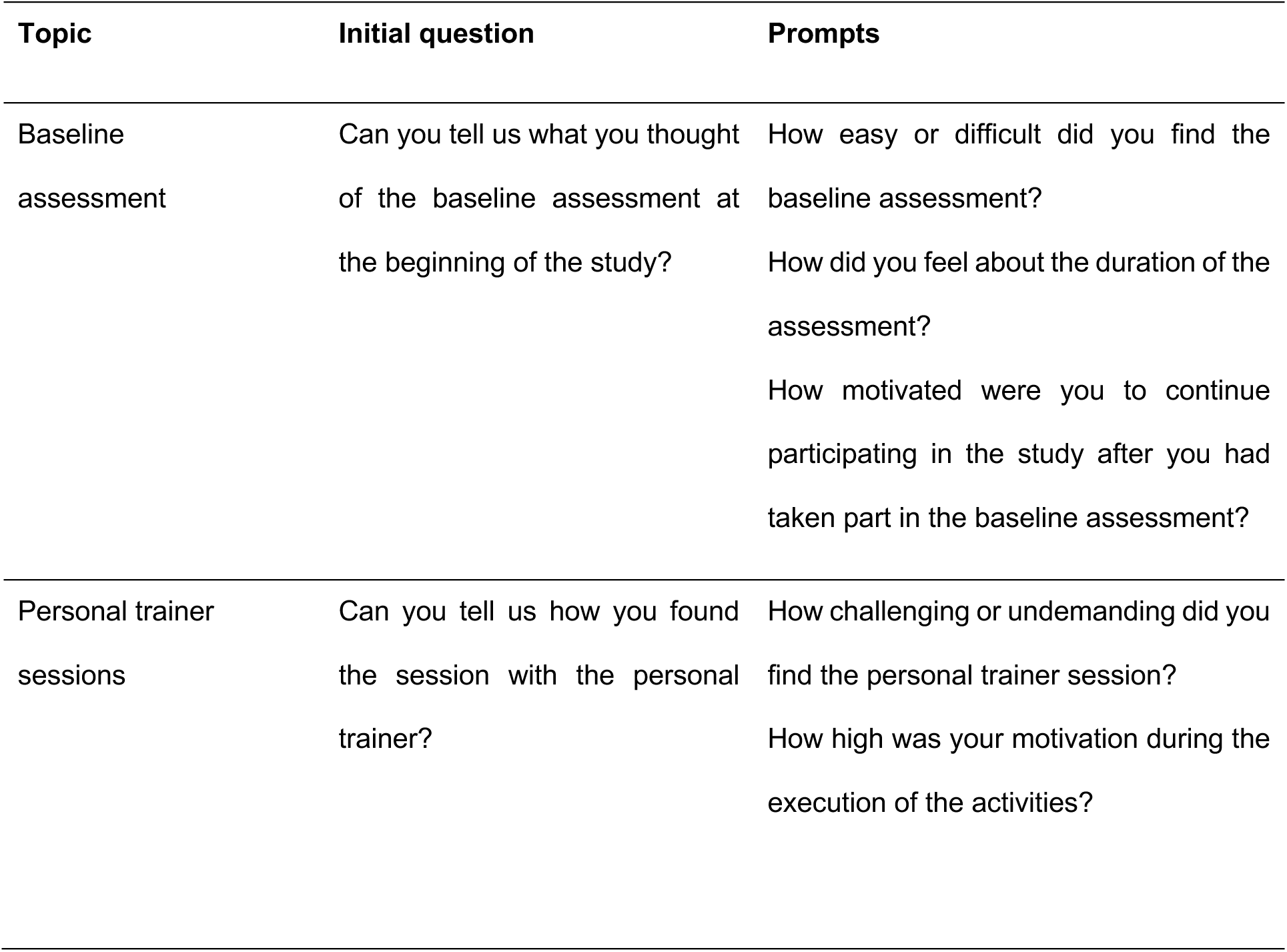
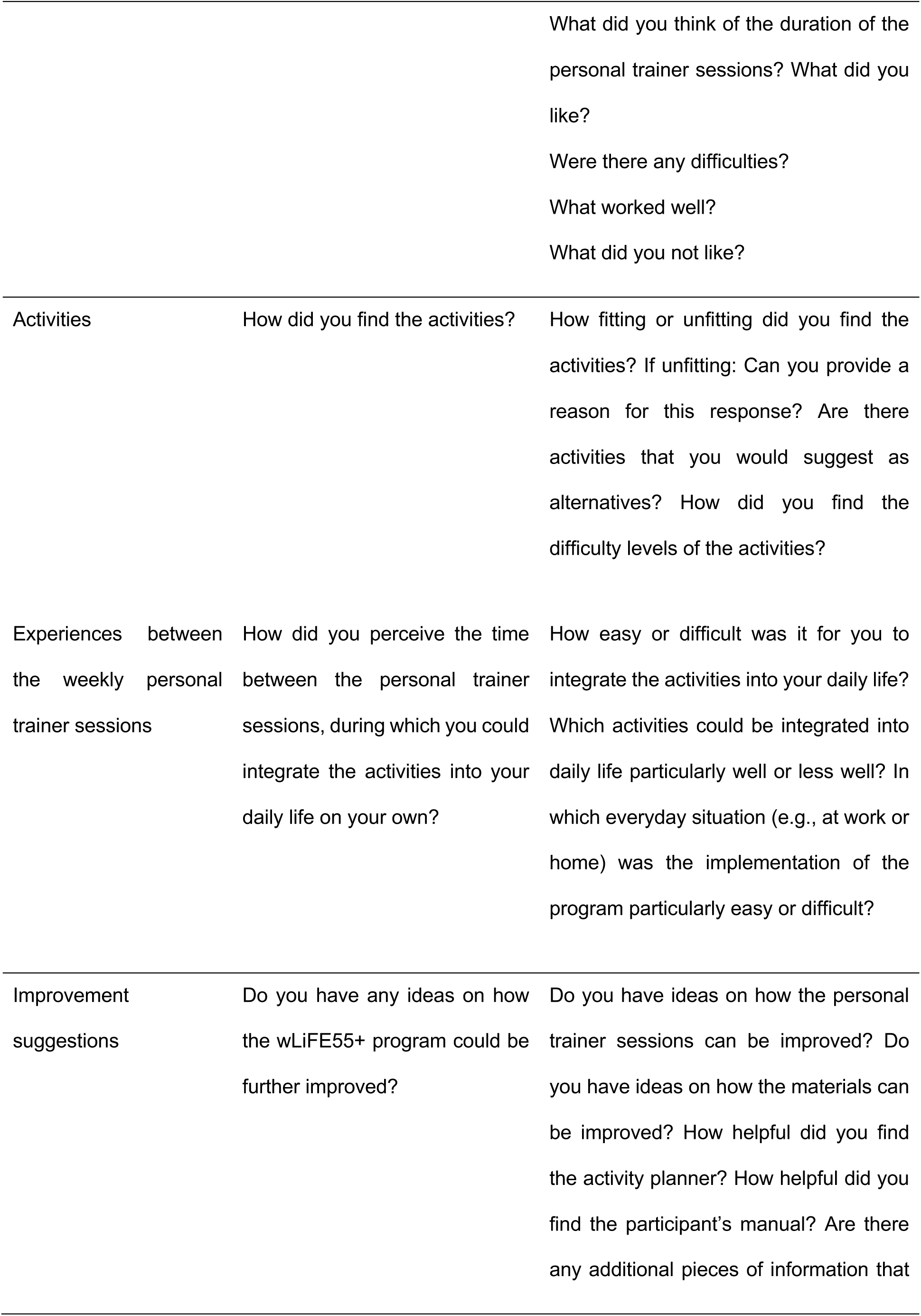
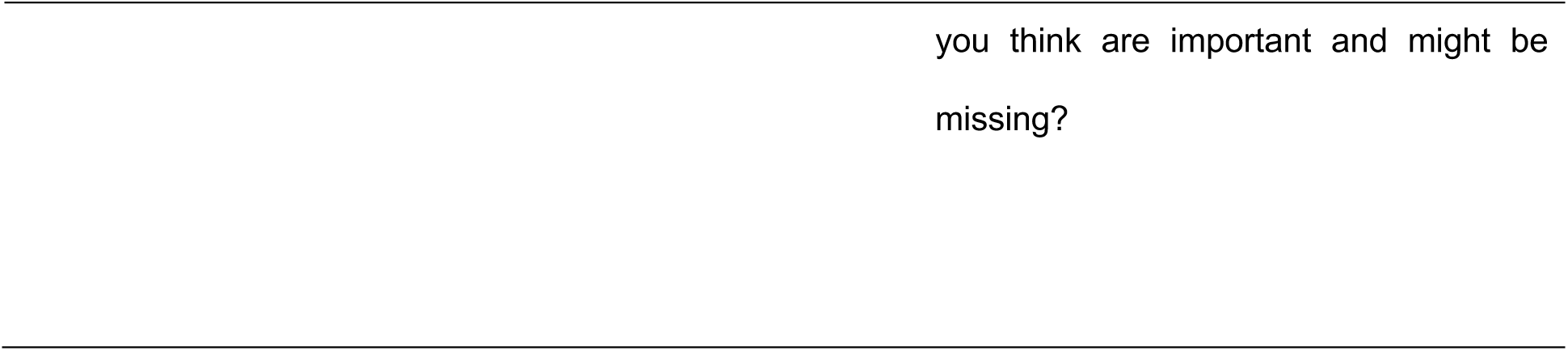
Semi-structured interview guide for wLiFE55+ focus groups.

| Topic | Initial question | Prompts |
| --- | --- | --- |
| Baseline assessment | Can you tell us what you thought of the baseline assessment at the beginning of the study? | How easy or difficult did you find the baseline assessment?<br>How did you feel about the duration of the assessment?<br>How motivated were you to continue participating in the study after you had taken part in the baseline assessment? |
| Personal trainer sessions | Can you tell us how you found the session with the personal trainer? | How challenging or undemanding did you find the personal trainer session?<br>How high was your motivation during the execution of the activities? |
|  |  | What did you think of the duration of the personal trainer sessions? What did you like?<br>Were there any difficulties?<br>What worked well?<br>What did you not like? |
| Activities | How did you find the activities? | How fitting or unfitting did you find the activities? If unfitting: Can you provide a reason for this response? Are there activities that you would suggest as alternatives? How did you find the difficulty levels of the activities? |
| Experiences between the weekly personal trainer sessions | How did you perceive the time between the personal trainer sessions, during which you could integrate the activities into your daily life on your own? | How easy or difficult was it for you to integrate the activities into your daily life?<br>Which activities could be integrated into daily life particularly well or less well? In which everyday situation (e.g., at work or home) was the implementation of the program particularly easy or difficult? |
| Improvement suggestions | Do you have any ideas on how the wLiFE55+ program could be further improved? | Do you have ideas on how the personal trainer sessions can be improved? Do you have ideas on how the materials can be improved? How helpful did you find the activity planner? How helpful did you find the participant's manual? Are there any additional pieces of information that |

**Table 4.**
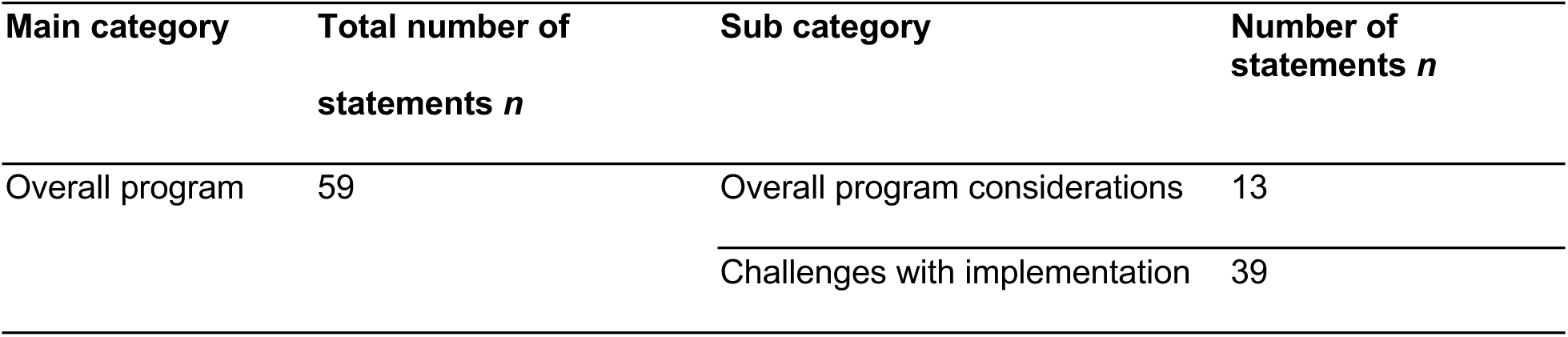
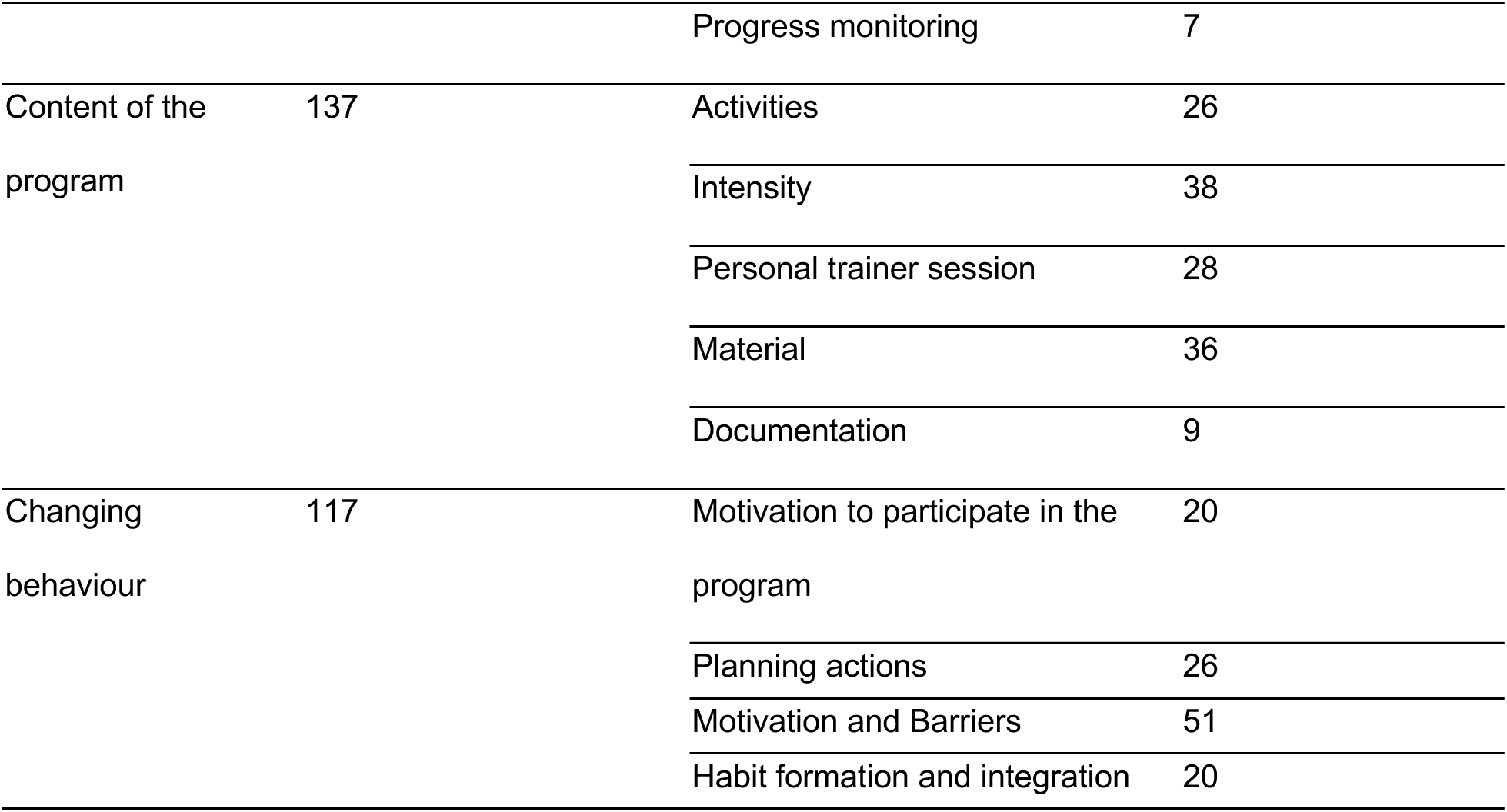
Overview number of statements.

| Main category | Total number of statements *n* | Sub category | Number of statements *n* |
| --- | --- | --- | --- |
| Overall program | 59 | Overall program considerations | 13 |
|  |  | Challenges with implementation | 39 |
|  |  | Progress monitoring | 7 |
| Content of the<br>program | 137 | Activities | 26 |
|  |  | Intensity | 38 |
|  |  | Personal trainer session | 28 |
|  |  | Material | 36 |
|  |  | Documentation | 9 |
| Changing<br>behaviour | 117 | Motivation to participate in the<br>program | 20 |
|  |  | Planning actions | 26 |
|  |  | Motivation and Barriers | 51 |
|  |  | Habit formation and integration | 20 |

### Data analysis

Data were analysed using qualitative content analysis following Mayring [33], which provides a systematic approach for examining implementation processes and health-related behaviours. A primarily deductive category system was developed based on previous LiFE studies [26, 34, 35] and the Theoretical Domains Framework (TDF) [36], which informed categories related to motivation, planning, capability and habit formation.

Initial categories were derived from a targeted literature search on qualitative evaluations of LiFE-based and workplace physical activity interventions (including aLiFE, gLiFE and iLiFE) [26, 34, 35] and were reviewed and refined by three researchers (DP, YR, MS), resulting in a structured coding guide with definitions and anchor examples (Supplementary Material 1).

All transcripts were imported into MAXQDA© and coded line by line using the smallest meaningful text segments as coding units, focusing exclusively on manifest content. During analysis, categories were iteratively adjusted to ensure conceptual fit, leading to a final structure of three overarching categories.

To ensure analytical rigour, two researchers initially coded the same material independently using the shared coding guide to check for a common understanding of the categories. Differences in coding were discussed and used to refine the coding framework. The revised framework was then applied in a second coding round with the involvement of a third researcher. Remaining differences were resolved through consensus discussions, a recognised quality-assurance strategy in qualitative content analysis [37, 38]. As the final codes were established through negotiated agreement rather than independent parallel coding, no intercoder reliability coefficient was calculated.

After coding, the material was condensed within categories and summarised, and representative quotations—including divergent perspectives—were selected to illustrate key themes in relation to the research questions.

### Researcher reflexivity and positionality

The research team consisted of sports science, occupational health and occupational safety experts. Two of the researchers (DP and YR) were involved in developing and implementing the wLiFE55+ pilot study, which could introduce interpretive bias. To mitigate this, an independent researcher (MS) moderated the focus groups.

## Results

### Results focus groups

A total of eight participants contributed 313 codable statements across the two focus groups. Three overarching themes emerged: (1) overall program experiences, (2) perceptions of program content, and (3) behaviour-change processes. Speaking time was balanced across participants (5–16%, M = 12.5%).

## 1. Overall Program

Participants generally described wLiFE55+ as meaningful and practical in a workplace context. The core idea of embedding strength and balance activities into everyday routines was consistently valued. Several participants noted that the program helped them recognise movement opportunities that “fit naturally into the day” (B3) and could be carried out without preparation or equipment. Even those who considered themselves physically active at baseline reported becoming more aware of intentionally integrating functional movements into their routine.

### 1.1 Overall program considerations

Participants suggested several ways to optimise organisational aspects of the program. A recurring theme was the desire for more time between personal trainer sessions, as the weekly interval felt too short to practise and consolidate new activities—especially when weekends intervened (B2). One participant summarised that “more practice time and habituation time in between” would have supported more stable learning (B3).

Another commonly mentioned issue was the need for a neutral, private room for trainer sessions. Participants working in shared offices reported discomfort practising balance or strength activities in front of colleagues. A separate space was described as more appropriate to avoid feeling watched or self-conscious (B5, B7).

Participants also expressed interest in expanding the program to include more upper-body activities, as exercises for the arms and trunk were perceived as underrepresented. They requested clearer explanations of what types of upper-body tasks could be incorporated into daily routines (B1, B4). Some further asked whether a follow-up study would be offered and expressed willingness to participate again (B1, B2).

### 1.2 Challenges with implementation

Challenges formed one of the most prominent subcategories and included structural, informational, and personal factors shaping early engagement. The baseline assessment was a major source of uncertainty: several participants were surprised by the strong emphasis on balance tests, especially the one-leg stand with closed eyes. Some described these tasks as “overwhelming at first” (B5), while others said they would have appreciated clearer information beforehand about what would be assessed and why (B1, B8). One participant even questioned whether they belonged to the target group because of the difficulty of the assessments (B6).

Participants also questioned whether the program was optimally matched to their demographic. Some felt that less active employees might benefit even more, noting that the group seemed relatively fit (B5, B6).

Workplace structure was another recurring barrier. Participants in open-plan offices reported feeling inhibited about practising visible exercises, such as tandem walking or stepping over objects. Unpredictable workdays—characterised by spontaneous meetings, sudden tasks, or periods of high workload—also disrupted activity plans. On stressful days, activities “became a nuisance” and were deprioritised despite positive intentions (B4).

Several participants further noted that the number of new activities introduced during the four weeks was high, leaving them feeling “overwhelmed” (B7) and unsure whether all tasks could realistically be integrated into everyday routines.

### 1.3 Progress monitoring and feedback

Participants expressed a clear interest in more differentiated feedback throughout the program. They wished to understand how their performance compared with age-matched reference values (B7) and found it motivating when trainers explicitly acknowledged improvements (B1). Many suggested the use of digital tools to enable automated feedback and easier progress tracking (B4). Some expressed curiosity about how progress would look after six to eight weeks, indicating a desire for a longer assessment horizon (B6).

## 2. Content of the Program

A total of 137 statements addressed participants’ experiences with specific program components, including activities, intensity, trainer support, materials, and documentation.

### 2.1 Activities

Participants expressed diverse views on the wLiFE55+ activities. Exercises that could be incorporated seamlessly into daily routines—such as one-leg stands while waiting at the printer, balance exercises during short pauses, or stair climbing between floors—were consistently described as intuitive, feasible, and natural (B3, B4). By contrast, activities requiring props or additional space were often viewed as impractical in an office setting (B2, B5). The tandem walk was repeatedly described as uncomfortable or unsuitable (B3, B4). Feedback on jumping activities was mixed: some found them energising (B2), whereas others felt they were unpleasant or too demanding (B7). Neuromotor tasks were sometimes described as surprising yet engaging, as they challenged participants in unexpected ways (B4).

Participants’ physical conditions strongly shaped perceptions; those with knee or joint discomfort adapted exercises accordingly (B5, B7).

### 2.2 Intensity

Intensity-related comments focused on both the baseline assessment and the weekly progression of activities. The baseline assessment was generally considered manageable but tiring due to its duration. The one-leg stand with closed eyes was repeatedly described as the most demanding component (B5, B7), and some noted the cognitive strain of completing multiple tests in one session.

During the intervention, perceptions of progression varied. Some participants appreciated the systematic increase in difficulty, finding it motivating. Others felt the progression was “too fast” (B3), particularly when they were still becoming familiar with earlier activities. Several mentioned the cognitive overload of learning and remembering numerous exercises within a short time frame (B7), which limited their ability to practise all tasks consistently.

### 2.3 Personal trainer sessions

Trainer sessions were consistently described as one of the strongest components of the program. Participants highlighted trainers’ empathy, responsiveness, and clarity of explanation. They valued receiving alternative strategies when difficulties arose and appreciated the encouragement to adapt exercises to personal ability and workplace constraints (B2, B3). Demonstrations were described as especially helpful for understanding correct execution (B7), and the respectful atmosphere reduced feelings of embarrassment or pressure.

A few participants noted that earlier, more explicit feedback in the first session would have reduced uncertainty (B1). Others emphasised how trainers actively sought tailored solutions when standard exercises were unsuitable (B2).

### 2.4 Materials and Documentation

Participants reported mixed experiences with the program materials. The wLiFE55+ manual was viewed as a helpful reference for recalling exercises once the trainer was absent, though participants recommended clearer illustrations and more detailed instructions.

The Activity Planner, however, was widely criticised. Participants described its structure as non-intuitive and difficult to integrate into daily routines. The need to navigate between multiple pages made it cumbersome (B4), and several admitted that they eventually stopped using it because documentation felt unnecessary for completing activities (B3, B7). Many strongly preferred a digital version that would reduce administrative burden and offer automated feedback (B4).

### 2.5 Documentation

Documentation requirements were often incompatible with participants’ work routines. Activities were frequently performed spontaneously, without the planner at hand (B3). Some found documentation redundant, noting that whether or not they recorded an activity had little influence on performing it (B7). This further reinforced participants’ preference for digital options.

## 3. Changing Behaviour

This category captures motivational dynamics, action planning, barriers, and habit formation.

### 3.1 Motivation to participate

Motivation at program start varied. Some participants described an immediate motivational increase after the baseline assessment, feeling encouraged to work on identified weaknesses (B2). Others maintained stable motivation throughout (B1). One participant expressed “cautious curiosity”, stating, “Let’s see how it turns out later” (B3).

A few participants indicated that because they already felt physically active, they perceived less urgency to adopt new routines, explaining they were “already on the road quite a bit” (B6). Nevertheless, many were curious about potential physical changes over time (B5).

### 3.2 Planning actions

Participants reported mixed experiences with the if–then planning strategy. Some found it a useful tool for initiating behaviour change (B2). Others struggled to generate meaningful situational cues during the trainer sessions and felt pressured to “come up with something spontaneously” (B2). Several later discovered that their chosen cues did not align well with their actual work routines (B1, B5). Over time, many participants independently adapted their activities to better match naturally occurring opportunities (B1, B3). Some requested more time during sessions to reflect on cues and test their applicability (B2, B4).

### 3.3 Motivation and barriers

This was the largest subcategory, illustrating the interplay between facilitators and barriers.

Early functional improvements—standing longer on one leg, increased stability on stairs, improved navigation in narrow spaces—served as strong motivators. Participants described balance improvements as occurring “very quickly, very well” (B3). Social experiences could also reinforce motivation: one participant used colleagues as a positive cue, reporting, “I used colleagues…that was really nice” (B4).

However, several barriers were identified. Negative experiences, such as stumbling, lowered confidence: “I stumbled around…it demotivated me a bit.” (B7). Rapid progression of difficulty also discouraged engagement: “Some of the steps were too big…which demotivated me” (B3). Social discomfort was a significant barrier. Participants avoided practising in visible areas due to fear of being observed or receiving negative comments, as reflected in the remark, “My colleague laughed in the background” (B7). Social dynamics were described as ambivalent—sometimes supportive, sometimes uncomfortable (B2).

Unpredictable work demands frequently interfered with activity plans. As one participant noted, “When tasks or stress suddenly get in the way…it felt more like a nuisance.” (B4).

### 3.4 Habit formation and integration

Habit formation varied widely across participants. Activities tied to recurring natural situations—particularly waiting times—were easiest to integrate and sometimes became semi-automatic. As one participant stated, “Wherever I wait, I automatically do one of the balance exercises” (B3).

Participants emphasised that forced or artificial cue–activity combinations rarely worked (B5). Many recognised early signs of routine development but felt that the four-week duration was insufficient for stable habit formation. Several estimated that six to eight weeks would be necessary (B6). Participants developed individualised strategies to flexibly integrate activities into unpredictable workdays, adjusting exercise selection and timing according to personal routines (B4, B6).

## Discussion

This qualitative evaluation provides an in-depth understanding of how older employees experienced wLiFE55+ and how lifestyle-integrated movements can be embedded into everyday workplace routines. Participants assessed the program as feasible and meaningful, particularly because the activities aligned with naturally occurring work tasks. These findings add to evidence from LiFE-based interventions, which consistently show that anchoring functional exercises in daily life enhances relevance, autonomy, and long-term adherence [25, 26, 34, 35, 39]. The present study is the first to extend this concept to an occupational context among older employees, providing new insights into how behaviour-change mechanisms manifest within organisational environments.

### Embedding lifestyle-integrated movement: alignment with work routines and behavioural theory

A central finding of this study was that perceived feasibility depended strongly on how well activities fitted into existing work routines. Activities that could be performed during naturally recurring situations—such as waiting times, short transitions, or routine movements—were described as intuitive and easy to adopt. In contrast, activities that required additional preparation, space, or visible performance were often perceived as awkward or impractical.

This finding directly reflects barriers consistently reported in workplace physical activity intervention research, including limited time, workflow disruptions, and misalignment with job demands [17, 40, 41]. Traditional workplace interventions are often perceived as add-ons rather than integral parts of the working day, which undermines sustained engagement. The present findings suggest that embedding movement into existing routines may overcome these structural barriers, supporting earlier claims that lifestyle-integrated approaches are particularly well suited for time-constrained populations [25, 39].

At the same time, participants’ experiences highlight that contextual fit is not solely determined by task structure but also by physical and social environments. Open-plan offices, shared workspaces and visibility to colleagues frequently inhibited activity performance. These findings underscore the importance of considering environmental opportunity, as conceptualised in the Capability–Opportunity–Motivation–Behaviour (COM-B) model [42], and align with previous workplace studies showing that organisational context can either enable or constrain health-promoting behaviours [19, 25, 39, 43].

### Behavioural mechanisms: autonomy, capability and early habit formation

Although behaviour change was not a primary aim of this four-week feasibility study, participants’ accounts provide insight into mechanisms relevant to engagement and early integration. The ability to adapt activities to personal routines and preferences was repeatedly emphasised and aligns with Self-Determination Theory, which highlights autonomy and perceived competence as key drivers of sustained behaviour [44].

Participants’ descriptions further suggest that early indicators of habit formation emerged when activities were linked to stable, frequently occurring cues. Waiting situations in particular appeared to provide reliable triggers that required minimal cognitive effort. This observation is consistent with habit formation theory, which posits that behavioural automatisation is facilitated by stable contexts and repeated cue–behaviour pairing [45].However, most participants felt that four weeks were insufficient for stable habit formation, estimating that at least six to eight weeks would be required. This perception aligns with empirical evidence suggesting that habit formation typically unfolds over longer timeframes [45, 46].

The if–then planning strategy was perceived as useful in principle but challenging to apply in dynamic work environments. Many participants reported difficulty identifying meaningful cues during training sessions, particularly because daily work routines were unpredictable. This finding reflects known limitations of implementation intention approaches in highly variable contexts and highlights the need for iterative refinement rather than one-time planning [47].

### Role of personal trainer support in implementation

Personal trainer support emerged as a key facilitator of engagement and perceived feasibility. Participants valued personalised instruction, adaptive problem-solving and empathetic communication. Trainers supported participants in tailoring activities to individual capabilities and workplace constraints, thereby enhancing perceived capability and confidence.

These findings are consistent with previous LiFE research, where guided personalisation was shown to support autonomous self-regulation and adherence [26, 34]. From an implementation perspective, trainer support appears particularly important in occupational settings, where heterogeneity in work tasks and environments necessitates flexible adaptation. However, participants also reported cognitive overload due to the rapid introduction of multiple activities within a short timeframe. This suggests that future implementations should prioritise fewer activities with slower progression to allow consolidation and reduce burden.

### Perceived benefits and motivational dynamics

Participants reported early improvements in balance and stability, which served as strong motivators for continued engagement. These mastery experiences enhanced self-efficacy and reinforced motivation, consistent with behaviour change theories emphasising the role of feedback and perceived competence [48, 49]. Conversely, negative experiences—such as stumbling or rapid increases in difficulty—were described as demotivating, underscoring the importance of carefully calibrated progression.

Social dynamics played an ambivalent role. While some participants experienced encouragement from colleagues, others felt self-conscious or discouraged when being observed. Similar ambivalence has been reported in prior workplace intervention studies, where social norms and peer reactions strongly influenced participation [43, 50]. These findings highlight that social context should be explicitly considered in workplace-integrated interventions.

### Implications for the refinement of workplace lifestyle-integrated interventions

The present findings yield several implications for the optimisation of wLiFE55+ and similar interventions. First, clearer preparatory communication is essential, particularly regarding assessment procedures and the focus on neuromotor activities, which were unexpected for many participants. Second, intervention duration and progression should be adjusted to better support learning and habit formation. Third, activity planning should be treated as an ongoing, adaptive process rather than a fixed component. Finally, environmental and social constraints within workplaces must be acknowledged and addressed to enhance feasibility. Although some participants expressed interest in digital documentation, digital tools should be considered optional rather than central, as adoption among older employees may be limited by digital literacy, perceived complexity, and additional cognitive burden.

These implications are consistent with implementation science frameworks emphasising flexibility, contextual adaptation and participant-centred design in complex interventions [40, 51].

## Conclusion

In summary, this qualitative evaluation demonstrates that workplace-integrated functional activities can be meaningfully adopted by older employees, while also revealing structural and organisational barriers that require refinement. Clearer preparatory communication, more gradual progression and longer intervention periods may strengthen feasibility and support more stable behavioural uptake. Future research should examine longer intervention durations and evaluate implementation and scale-up in larger and more diverse organisational settings to support sustainable workplace integration and active ageing in predominantly sedentary work environments.

## Strength and Limitation

A key strength of this study is the balanced participation within both focus groups, which ensured that no single participant dominated the discussion and allowed for a broad range of perspectives [52]. The open and reflective group atmosphere, including the willingness to discuss challenges, suggests limited social desirability bias and supports the credibility of the findings. Methodological rigour was further enhanced by the systematic application of qualitative content analysis following Mayring’s framework and by conducting two small, homogeneous focus groups, which are well suited to generating rich, in-depth data [33]. The use of an independent facilitator to conduct the focus groups reduced the risk of subjective bias and researcher influence [26].

Several limitations should be acknowledged. The four-week intervention period was appropriate for feasibility testing but likely insufficient for full integration and stable habit formation, particularly given the number of activities introduced. The qualitative sample was small and drawn from a single pilot trial, which limits transferability to other workplace settings and occupational groups. Methodologically, the primarily deductive coding approach may have constrained the emergence of unanticipated themes, and no formal intercoder reliability coefficient was calculated, as coding decisions were resolved through consensus. Finally, non-verbal communication was not analysed, and social desirability effects—common in focus group research—cannot be fully excluded.

## List of abbreviations

COM-B: Capability–Opportunity–Motivation–Behaviour model
PA: physical activity
WPAI: workplace physical activity intervention
wLAT55+: work LiFE55+ Assessment Tool
wLiFE55+: work Lifestyle-integrated Functional Exercise Program for employees over 55 years

## Declarations

### Ethical approval and consent to participate

All participants provided written informed consent prior to their involvement in the study. The study protocol was reviewed and approved by the Ethics Committee of the University of Konstanz, under the ethics approval number IRB23KN07-005w.

### Consent for publication

All the participants who provided their case reports provided consent for publication.

### Availability of data and materials

All data generated or analyzed during this study are included in this published article and its supplementary information files.

### Authors’ contributions

YR developed the concept and design of the study and was responsible for study management as well as the statistical analysis and interpretation of the quantitative data. DP was responsible for writing the manuscript and was involved in study management, qualitative data collection, and analysis, including the development of the interview guide, transcription, and evaluation of the focus groups.

MS contributed to obtaining funding, the development of the study concept and design, the implementation and moderation of the focus groups, and the interpretation of the data.

All authors contributed to the interpretation of the data, the writing of the article and the final approval of the version to be published.

## Data Availability

All data produced in the present work are contained in the manuscript.

## Acknowledgments

The authors cordially thank the wLiFE55+ participants for making this research possible by taking part in the study.

The authors further thank Alina Boden (AB) (University of Konstanz) for contribution in supporting the coding of participant statements.

## Authors’ details (optional)

Not applicable

